# COVIDReady2 Study Protocol: Cross-sectional Survey of Medical Student Volunteering and Education During the COVID-19 Pandemic in the United Kingdom

**DOI:** 10.1101/2021.02.24.21252103

**Authors:** Matthew H V Byrne, James Ashcroft, Laith Alexander, Jonathan C M Wan, Anmol Arora, Megan E L Brown, Anna Harvey, Andrew Clelland, Nicholas Schindler, Cecilia Brassett, Rachel Allan on behalf of the MedEd Collaborative

**Author notes:** Corresponding author: Name: Matthew Byrne, Email address, Institution address: Churchill Hospital, Old Rd, Headington, Oxford, OX3 7LE, UK. Collaborating authors are listed in Appendix A. Funding declarations: None. Conflicts of interests: None. Study type: Protocol. Ethics: Ethical approval for this study was obtained from the University of Oxford Medical Science Interdivisional Research Ethics Committee (Reference: R74003/RE001). Contributions: MHVB, JA, JCMW, LA were responsible for conceptualisation. All authors were responsible for writing the first draft. All authors and collaborative authors under the group authorship ‘MedEd Collaborative’ were responsible for designing the survey. All authors and collaborative authors under the group authorship ‘MedEd Collaborative’ were responsible for revisions. RA was responsible for supervision. MedEd Collaborative Twitter: @MedEdCollab.

## Abstract

**Background and objectives:** Covid-19 has led to global disruption of healthcare. Many students volunteered to provide clinical support. Volunteering to work in a clinical capacity was a unique medical education opportunity; however, it is unknown whether this was a positive learning experience or which volunteering roles were of most benefit to students.

**Methods and Design:** The COVIDReady2 study is a national cross-sectional study of all medical students at UK medical schools. The primary outcome is to explore the experiences of medical students who volunteered during the pandemic in comparison to those who did not. We will compare responses to determine the educational benefit and issues they faced. In addition to quantitative analysis, thematic analysis will be used to identify themes in qualitative responses.

**Discussion:** There is a growing body of evidence to suggest that service roles have potential to enhance medical education; yet, there is a shortage of studies able to offer practical advice for how these roles may be incorporated in future medical education. We anticipate that this study will help to identify volunteer structures that have been beneficial for students, so that similar infrastructures can be used in the future, and help inform medical education in a non-pandemic setting.

## INTRODUCTION

### Background

The coronavirus disease 2019 (Covid-19) pandemic has led to global disruption of healthcare services and medical education. In March 2020, the UK government announced that medical students were permitted to provide assistance during the pandemic.^1^ Almost 7000 final year students obtained early provisional registration from the General Medical Council (GMC), filling 4662 new foundation interim year 1 (FiY1) doctor posts between April to July 2020. Students in other years volunteered to work in a wide range of roles to provide clinical support.^2–5^ Similar initiatives have been organised internationally.^6–8^

The UK Medical Schools Council advised against volunteering if it would disrupt a student’s studies.^9^ However, as many medical students had their medical studies and clinical placements interrupted, clinical volunteering work provided a unique medical educational experience to continue clinical skills development and gain additional skills, albeit in an unusual fashion. A survey of 440 final year medical students from 32 medical schools across the UK near the start of the pandemic indicated that students generally agreed that changing conventional placements due to the pandemic was necessary, but that they also felt less prepared for beginning their work as a doctor.^10^

Our first COVIDReady study evaluated responses from 1145 students at 36/42 UK medical schools from 2^nd^ May to 15^th^ June 2020 (University of Cambridge Psychology Research Ethics Committee reference PRE.2020.040).^11^ Of these, 82.7% of students were willing to volunteer. The majority of students (86.6%) felt that volunteering would benefit their education, and this was the strongest predictor of willingness to volunteer on multiple regression analysis (estimate=0.35±0.03, adjusted P<0.001). Thematic analysis of free text responses of anticipated issues when volunteering identified five themes: Education; Finances and Logistics; Professional Practice; Pressure to Volunteer; and Safety.^11^ These themes broadly concur with the three main motivators towards joining the workforce early as identified by a GMC survey of 1448 final year students: learning gain, altruistic reasons and financial gain. The GMC also reported that a notable motivating factor was that volunteering would allow students to leave the house and socialize during lockdown conditions in which electives and holidays were cancelled.^3^ Overall, the body of literature exploring student attitudes towards Covid-19 volunteering is maturing, but there is a notable shortage of studies aimed at students after they have actually participated in these roles.^12–14^ One local study in Hamburg, Germany found that a majority of final year students (n=40) working in hospitals during the pandemic felt that their help was not appreciated, that they were inadequately protected when treating Covid-19 patients, and that they did not acquire new skills.^12^ Interestingly, in the same study, students in younger years who were volunteering (n=17) generally felt appreciated, adequately protected and valued the learning experience. These findings suggest a complex interplay between the design of volunteering programmes during the pandemic and outcomes for students. Exploring these outcomes at a national scale remains a priority area of medical education research.

### Rationale for this study

Early research has explored the uptake of volunteering opportunities by students but has not explored whether volunteering was a positive learning experience. This broadly reflects literature from previous pandemics and other disasters.^15,16^ The COVIDReady2 study aims to build on previous work by assessing the educational benefit of volunteering and exploring the issues students faced while volunteering during the pandemic. We aim to answer the research questions: *‘Was volunteering (in a paid or unpaid clinical capacity) during the pandemic a positive experience for medical students and what were the reasons for this? What issues did volunteer and non-volunteer medical students encounter with volunteering during the pandemic?’* We anticipate that this study will help to identify volunteer structures that have been beneficial for students, both so that similar infrastructures can be used in the future and to inform the structure of the non-pandemic clinical placements.

### Study objectives

#### Primary objective

To explore the effect of volunteering during the pandemic on the medical education of students who volunteered, in comparison to those who did not volunteer.

#### Secondary objectives

To identify:

- Whether students would be willing to assume similar roles in a non-pandemic setting
- If students found the experience more or less beneficial than traditional hospital placements and the reasons for this
- Students’ perceived benefits and disadvantages of volunteering
- Difference in perceived preparedness between students who did and did not volunteer for FY1 and the next academic year

To explore:

- Issues associated with volunteering, including safety concerns
- Issues with role and competence

### Ethics

Ethical approval for this study was obtained from the University of Oxford Interdivisional Medical Sciences Research Ethics Committee (Reference: R74003/RE001).

## METHODS

### Study Design

The COVIDReady2 study is a cross-sectional study of students and foundation year one (FY1) doctors who were medical students at UK medical schools between March 2020-June 2021 during the COVID-19 pandemic of 2020. The study will consist of an online survey disseminated to medical students and foundation doctors over a six-week period in Spring 2021.

### Study population

Students and current FY1 doctors who were enrolled at a UK medical school at the start of the first UK lockdown for the COVID-19 pandemic (23rd March 2020), regardless of whether or not they volunteered during the pandemic.

### Inclusion criteria

- Medical students at UK medical schools identified by the Medical Schools Council or FY1 doctors, regardless of whether or not they volunteered during the pandemic.
- Students and FY1 doctors must have been enrolled at a UK medical school at the start of the first UK lockdown for the COVID-19 pandemic (23^rd^ March 2020).
- Participants must be aged 18 or older.

### Exclusion criteria

- Students commencing their first year of medical school in the academic year 2020-2021.

### Survey development

The survey was developed from existing literature following a systematic review of studies assessing willingness to volunteer during pandemics and disasters, the first COVIDReady study, and consultations with medical education specialists and medical students. A pilot study of 20 students was performed to ensure questions were unambiguous and establish face validity. The study was reviewed and approved by the UK Medical Schools Council Education Lead Advisory Group.

The survey will be hosted on Qualtrics XM (USA).^17^ The survey will be preceded by a description of the study, the research question, the contact details of the primary investigator, and a consent form. Data will be held on a secure server and email addresses, provided for optional follow up, deleted in two years. Once starting the survey, participants can withdraw consent by closing the browser and no data will be collected. As no identifiable data is collected participants will not be able to withdraw consent following completion of the survey, unless they have chosen to provide an email address at the end of the survey. The survey can be found in Appendix B.

## Data collection

All medical schools will be contacted via their medical school office general enquiries email identified through a list provided by the Medical Schools Council.^18^ This email will ask the medical schools to distribute a link to the survey to medical students and recently graduated medical students who are now working as FY1 doctors. We aim to recruit medical students from each UK medical school and FY1 doctors from each foundation school to help distribute the survey via social media. Messages will be posted at a maximal rate of once per week to prevent excessive messaging to students. After completion of the survey, participants are asked to share the survey with three other medical students to recruit additionally participants via a snowball approach.

### Sample size calculation and rationale

A sample size calculation was performed, which showed that a total of 630 respondents are required to identify a significant 0.5 point (out of a 5-point Likert scale) increase in confidence or agreement with the statement ‘I believe the period during the first lockdown (23 march 2020 to 4 July 2020) has benefited my medical education / career’, considering stratification by year group.

This power calculation was performed with an alpha of 0.05 and a power of 80%. Based on the results from the first COVIDReady study, the standard deviations of the anticipated Likert score means were conservatively set to 1 (out of 5), the ratio of students volunteering/not-volunteering was estimated at 1:1, and the year group distribution of students was assumed to be equal.

### Data analysis plan

STROBE guidelines for cross-sectional studies will be followed.^19^ Data will be presented as descriptive statistics and comparisons between volunteer and non-volunteer groups. For questions where qualitative responses are collected, we will conduct a thematic analysis using Braun and Clarke’s reflexive approach to thematic analysis to code, sort, and analyse data.^20^ Medical school names will be anonymised.

### Expected outputs

We anticipate that the results of this study will be published on a pre-print server and subsequently in a peer-reviewed medical or scientific journal. We will submit this project to national and international conferences. Individuals involved in the recruitment of participants will be listed as collaborators under the group authorship: “MedEd Collaborative”.

## DISCUSSION

Disruption caused to medical education by the ongoing Covid-19 pandemic has been well described; however, subsequent sequalae such as volunteering and early graduation remain under-discussed.^21–23^ One way volunteering may benefit students is through increased service-based learning, a method of teaching wherein students perform and reflect on roles which intersect with their academic curriculum while also addressing community needs.^24^

Arguments for adding service roles to curricula are grounded in educational theory, but due to medical school curricula constraints this has not been possible to achieve formally to date. The medical profession relies on experiential learning to refine situational judgement.^25,26^ Lave and Wenger’s Communities of Practice (CoP) theory posits that learning is driven by novices working alongside experts and through the process of legitimate peripheral participation they move towards becoming experts themselves.^27–29^ The theory was not intended to be a prescriptive model but rather a framework which may guide how to maximise knowledge sharing.^30,31^ Gaining experience as part of a clinical team and openly reflecting on it in informal settings can allow students to acquire tacit knowledge and personal growth which formal teaching cannot achieve.^27^ In this way, even if service-learning roles do not directly offer knowledge-based teaching, they may improve self-confidence, analytical abilities and communication skills.^32^ The argument that integrated service roles may improve interpersonal skills is also supported by research concerning the educational principle of continuity, which suggests that spending extended periods of time as part of a medical team and following patient care throughout illness episodes may be beneficial.^33^ However, it could also be contended that if service roles were not designed and integrated with the principle of continuity in mind, they may further fragment schedules and disrupt existing continuity, including examination timelines. In this way, it is possible that the effects of the pandemic and volunteering may have disrupted existing continuity within medical education.In the future, if such roles were integrated alongside conventional placements rather than disrupting them, they may introduce a fresh degree of continuity. This continuity is key to longitudinal integrated clerkships, a role in which students stay in the same place, with the same group and the same clinical team for extended periods of time.^34^

Differences between conventional clinical placements and service roles are subtle, since they could involve students being in identical clinical environments but performing different activities. Whilst ‘conventional’ placements are heavily focused around students extracting learning potential from clinical environments, service roles involve balancing the objectives of additional stakeholders, such as the service provider.^35^ O’Byrne et al. (2020) suggested that, whilst conventional placements can provide a stable learning environment, they may not provide students with the confidence and skills that might be required during a pandemic.^36^ Conventional placements may be designed to focus on students maximising learning experience, and moving too far towards service provision may too be detrimental.^32^ A concern that has been raised about service learning pertains to the heterogeneity between different placements, both in terms of the quality of the learning experiences and the grading of these learning experiences.^37^ By contrast, standard clinical placements often revolve around predefined learning objectives which correspond to standardised assessment criteria.^38^ This could be attributed to the principal of constructive alignment, which has been widely adopted in medical education in order to ensure that teaching and assessment are aligned with required learning objectives.^39,40^

Service-based learning is not equivalent to conventional volunteering, such as the type which is likely to have occurred during this pandemic. Service-based learning incorporates structured volunteer-based activities with pre-defined learning objectives and, crucially, reflective practice.^32^ It has been postulated that whilst ‘service-learning’ may offer benefits to students, ‘community service’ may hinder their progress.^32^ On this basis, it is plausible to argue that pandemic voluntary experiences could have been detrimental to medical education. The volunteering that may have occurred during the pandemic was likely to have been unstructured and independent of medical school learning objectives. Such volunteering is commendable, but may offer only incidental or occasional learning opportunities, rather than the clearly defined outcomes of conventional placements. During any future waves of COVID-19, as well as other potential disasters, it may be more beneficial to employ principles of constructive alignment by formalising volunteering roles as medical education opportunities, with defined outcomes and reflection.^40,41^ Prior to the pandemic, there were suggestions for increased provision of service-based learning beyond the clinical experience achieved through traditional clinical placements within UK medical curricula.^32^ In the UK, these suggestions have been in the form of apprenticeship style final-years.^42–44^ Though there has been some study of apprenticeships, there has been little empirical research able to recommend specific program structures and implementation steps for service-based learning, especially in earlier years of the medical curriculum.^24,45^ However, early efforts to introduce service roles have shown promising results.^46^

The year of medical school in which such programmes are implemented may be an independent factor in determining the success of the programme. For example, Elam et al (2003) suggested that incorporating service-based learning early in curricula may help in developing a community-based perspective towards clinical practice.^47^ Working in voluntary roles under the supervision of nurses and allied healthcare professionals teaches the importance of interprofessional collaboration in delivering care, an early understanding of which can reap dividends in future years of medical training.^48^ Importantly, there is only a limited body of evidence exploring practical considerations of volunteering roles. The early literature does generally concur that matching activities to learning goals and organising reflective learning and assessments can contribute to successful service learning structures.^32^ This study aims to build on that body of literature by identifying volunteering structures which students found useful during the pandemic, as well as those that they did not.

The current study presents a unique opportunity for research whereby we are able to compare attitudes and perceived outcomes of students who volunteered with those who did not on a large scale. This will help to inform volunteering infrastructure during future waves of COVID-19, as well as informing the structure of current clinical placements and apprenticeships. Our findings may also have implications for re-structuring overseas medical electives where students volunteer without clearly defined curricular or learning objectives.

Our first COVIDReady study sought to explore the willingness to volunteering during the Covid-19 pandemic; COVIDReady2 now seeks to explore the educational impact of volunteering. In the short term, our findings may inform infrastructures for medical student volunteering in future waves of the Covid-19 pandemic. In the long term, our findings may inform the structure of clinical placements, electives, and apprenticeships. We anticipate future studies may explore the effects of volunteering on academic performance and career progression.

## Data Availability

Not applicable. All relevant information is contained within the manuscript.

## DECLARATIONS

### Ethics approval and consent to participate

Ethical approval for this study was obtained from the University of Oxford Medical Science Interdivisional Research Ethics Committee (Reference: R74003/RE001).

### Consent for publication

Not applicable

### Authors’ contributions

MHVB, JA, JCMW, LA were responsible for conceptualisation. All authors were responsible for writing the first draft. All authors and collaborative authors under the group authorship ‘MedEd Collaborative’ were responsible for designing the survey. All authors and collaborative authors under the group authorship ‘MedEd Collaborative’ were responsible for revisions. RA was responsible for supervision.

## Acknowledgements

We would like to thank the UK Medical Schools Council, the Royal Society of Medicine Student Members Group, IncisionUK, and the ADAPT Consortium for their support in developing this study.

## Availability of data and materials

Not applicable

## APPENDIX A MedEd Collaborative Authorship (all co-authors PubMed citable)

### Writing Group

Matthew H V Byrne, James Ashcroft, Laith Alexander, Jonathan C M Wan, Anmol Arora, Megan E L Brown, Anna Harvey, Andrew Clelland, Nicholas Schindler, Cecilia Brassett, Rachel Allan (Senior Author)

### Survey development group

Bryan Burford, Gillian Vance, Vigneshwar Raj, Soham Bandyopadhyay, Catherine Dominic, Siena Hayes, Aleksander Dawidziuk, Florence Kinder, Sanskrithi Sravanam, Michal Kawka, Adam Vaughan, Oliver P Devine, Aqua Asif, Jasper Mogg. All members of the writing group also contributed to survey development.

## APPENDIX B SURVEY

*Compulsory (only three questions)

| **Question** | **Answer options** |
| --- | --- |
| **1. Demographics** |  |
| Did you volunteer or work in a clinical capacity during the pandemic?* | Yes/No |
| Year at medical school when the COVID-19 pandemic started (not counting any intercalated years)* | DROP DOWN 1-5, intercalation year 3, intercalation year 4 |
| Are you on a four-year graduate entry medical course?* | Yes/No |
| Medical school | DROP DOWN of all UK medical schools & prefer not to say |
| Age (at time of volunteering) | NUMERICAL, limit 16-70 years old |
| Gender | MULTIPLE CHOICE Male, female, non-binary, prefer not to say, other [free text] |
| Ethnicity | MULTIPLE CHOICE<br>White - British<br>White - Irish<br>White - Gypsy or Irish Traveller<br>White - Any other white background<br>Mixed/Multiple ethnic groups - White and Black Caribbean<br>Mixed/Multiple ethnic groups - White and Black African<br>Mixed/Multiple ethnic groups - White and Asian<br>Mixed/Multiple ethnic groups - Any other mixed/multiple ethnic background<br>Asian/Asian British - Indian<br>Asian/Asian British - Pakistani<br>Asian/Asian British - Bangladeshi<br>Asian/Asian British - Chinese<br>Asian/Asian British - Any other Asian/Asian<br>British Background<br>Black/African/Caribbean/Black British - African<br>Black/African/Caribbean/Black British - Caribbean<br>Black/African/Caribbean/Black British - Any other<br>Black/African/Caribbean/Black British<br>Other ethnic group - Arab<br>OTHER (FREE TEXT)<br>Prefer not to say |
| **QUESTIONS ONLY IF THEY VOLUNTEERED** |  |
| **Volunteer role** |  |
| Which deanery did you volunteer in? | DROP DOWN<br>East Anglia<br>Essex, Bedfordshire & Hertfordshire (EBH)<br>Leicester, Northamptonshire & Rutland (LNR)<br>London & KSS: (North Central and East London; North West London; South Thames)<br>North West of England<br>Northern<br>Northern Ireland |
|  | Oxford<br>Peninsula<br>Scotland<br>Severn<br>Trent<br>Wales<br>Wessex<br>West Midlands (Central, North & South)<br>Yorkshire & Humber |
| Is your medical school in this deanery? | Yes/No |
| How many miles away from your home did your volunteering role take place? | 0<br>1-10<br>11-20<br>21-30<br>31-40<br>41-50<br>50+ |
| When did you start volunteering or working? | DATE |
| How many weeks did you volunteer or work for? | NUMERICAL VALUE |
| Which months did you volunteer in? | CHECKBOX<br>March 2020<br>April 2020<br>May 2020<br>June 2020<br>July 2020<br>August 2020<br>September 2020<br>October 2020<br>November 2020<br>December 2020<br>January 2021<br>February 2021<br>March 2021<br>April 2021<br>May 2021 |
| On average how many hours a week did you volunteer or work for? | MULTIPLE CHOICE<br>0<br>1-5<br>6-10<br>11-15<br>16-20<br>21-25<br>26-30<br>31-35<br>36-40<br>41-45<br>46-50<br>50+ |
| Was this a paid role? | Yes/No |
| Where did you volunteer or work? | Hospital<br>GP<br>OTHER (FREE TEXT) |
| What was the main role you had when volunteering or working? | MULTIPLE CHOICE<br>Administrative role (e.g. ward clerk, secretary, receptionist, 111 caller)<br>Allied healthcare professional/technician |
|  | Carer/care assistant (in hospital or care home)<br>Dentist<br>Doctor (interim foundation or equivalent)<br>First responder/first aider<br>Healthcare assistant<br>Healthcare scientist (e.g. hospital laboratory, clinical or laboratory research)<br>Nurse<br>Paramedic/emergency medical technician<br>Pharmacist<br>Pharmacy assistant<br>Phlebotomist<br>Doctor's assistant<br>Patient liaison<br>Primary care (GP) role eg: summariser, HCA, QI/Audit<br>Vaccinator<br>OTHER (FREE TEXT) |
| Did you hold this role prior to the pandemic? | Yes/No |
| Please describe what this role involved | FREE TEXT |
| **Opinions** |  |
| How did volunteering benefit your learning? (both as a medical student and preparation to practice as a Foundation Year 1 doctor) | FREE TEXT |
| How did volunteering disadvantage your learning? (both as a medical student and preparation to practice as a Foundation Year 1 doctor) | FREE TEXT |
| I would be willing to perform the same role in a non-pandemic setting as a formal part of my medical degree programme | LIKERT |
| I found my role as a volunteer more useful for my learning, than my role as a medical student on a clinical placement in a similar setting | LIKERT |
| Why did you find the role as a volunteer more or less useful than your role as a medical student on a clinical placement in a similar setting? | FREE TEXT |
| How has your volunteering experience affected your approach to patients? | FREE TEXT |
| Has your volunteering experience changed your view of how the NHS or other healthcare facilities work and if so how? | FREE TEXT |
| Were any opportunities provided for reflective practice? | Yes/No |
| **IF YES TO REFLECTIVE PRACTICE** |  |
| What opportunities were made for reflective practice? | CHECKBOX<br>Portfolio logs<br>Diaries<br>In-person reflective discussions<br>OTHER (FREE TEXT) |
| I found the reflective practice provisions useful. | LIKERT |
| **IF NO TO REFLECTIVE PRACTICE** |  |
| I think reflective practice provisions would have been useful. | LIKERT |
| **Issues** |  |
| What issues did you encounter while volunteering during the coronavirus pandemic? | FREE TEXT |
| Did you do on-calls? | MULTIPLE CHOICE<br>None<br>Day<br>Night<br>Day and Night |
| Did you work with patients who were COVID-19 positive? | Yes/No |
| I understood my role and the specific tasks I was expected to undertake | LIKERT |
| I felt competent to perform my role. | LIKERT |
| I felt other staff were aware of my role and the specific tasks I was expected to undertake | LIKERT |
| I was asked to perform skills outside my competence. | LIKERT |
| I felt confident saying no to tasks I was not adequately prepared for. | LIKERT |
| I felt patient safety was compromised while volunteering. | LIKERT |
| I received adequate supervision at work. | LIKERT |
| Did you have a named supervisor? | MULTIPLE CHOICE<br>Yes - clinical and educational supervisor<br>Yes - clinical supervisor only<br>Yes - educational supervisor only<br>No |
| I received a contract | MULTIPLE CHOICE<br>Yes - prior to work<br>Yes - after starting work<br>No |
| I was told who I could speak to if I had personal, health, or mental health problems while at work | Yes/No |
| What additional training did you receive for your role? (Please tick all that apply) | CHECKLIST<br>None<br>Induction<br>How to report clinical incidents (e.g. Datix)<br>First aid training<br>Basic life support (BLS) training<br>Basic infection control training (e.g. handwashing)<br>Surgical scrubbing and gowning<br>Infection control training with personal protective equipment (e.g. respirator masks, donning and doffing of goggles and garments)<br>Pandemic influenza teaching<br>OTHER (FREE TEXT) |
| Did you test positive for COVID-19 while volunteering? | Yes - swab test positive<br>Yes - swab test negative, but confirmed by health professional<br>Yes - suspected due to symptoms (e.g. fever, cough, shortness of breath, loss of smell)<br>No |
| **QUESTIONS ONLY IF THEY DIDN'T VOLUNTEER** |  |
| What were reasons that you did not volunteer? | FREE TEXT |
| Have you had any negative experiences by not volunteering? | Yes/No |
| If you did have negative experiences by not volunteering what were they? | FREE TEXT |
| **QUESTIONS FOR ALL** |  |
| I would be willing to volunteer to work in the future if the need arose. | LIKERT |
| I believe the period from the start of the first lockdown (23 March 2020) until now has benefited my medical education / career. | LIKERT |
| I believe the period during the first lockdown (23 March 2020 to 4 July 2020) has benefited my medical education / career. | LIKERT |
| The skills I have developed during the first lockdown (23 March 2020 to 4 July 2020) have prepared me for FY1 | LIKERT |
| The skills I have developed during the first lockdown (23 March 2020 to 4 July 2020) have prepared me for the 2020-2021 academic year | LIKERT |
| How have these skills prepared you for FY1 or the 2020-2021 academic year? | FREE TEXT |
| I feel assistantships (final year clinical placements) should be replaced with formal roles within a clinical team | LIKERT |
| What are your thoughts on replacing assistantships (final year clinical placements) with formal roles within a team (paid or unpaid)? | FREE TEXT |
| How many times did you perform the following skills during the first lockdown (23 March 2020 to 4 July 2020): | Venepuncture<br>Cannulation<br>Prescription of medications<br>History or examination of patient<br>Organise investigations for patients (e.g. blood tests, x rays etc)<br>Formulate management plans<br>Prescribe fluids<br>Measure observations/vital signs<br>Communicate information to patients<br>Communicate information to other healthcare professionals<br>Administrative medical duties (e.g. documentation, discharge letters etc)<br>Complete a chronic disease review<br>Communicate with patients as part of a medication review or audit<br>MULTIPLE CHOICE for each skill<br>None<br>Observed<br>Assisted<br>Performed 1-5 times (independently)<br>Performed 6-10 times (independently)<br>Performed 10+ times (independently) |
| What support did you receive from your medical school to facilitate volunteering? | FREE TEXT |
| Follow up study |  |
| Please enter your email address if you would be willing to be contacted for a future study.<br>We may conduct a follow up study to understand in greater depth responses from this study. This would either be by a focus group discussion with a researcher and other survey participants, by a semi-structured interview with a researcher, or an additional survey. By entering your email address it does not commit you to participating in a future study.<br>Your email address will be separated from the data entered before the data is analysed, will not be shared outside the data collection team, and will be deleted in two years time. | EMAIL |

